# BNT162b2 and mRNA-1273 COVID-19 vaccine effectiveness against the Delta (B.1.617.2) variant in Qatar

**DOI:** 10.1101/2021.08.11.21261885

**Authors:** Patrick Tang, Mohammad R. Hasan, Hiam Chemaitelly, Hadi M. Yassine, Fatiha M. Benslimane, Hebah A. Al Khatib, Sawsan AlMukdad, Peter Coyle, Houssein H. Ayoub, Zaina Al Kanaani, Einas Al Kuwari, Andrew Jeremijenko, Anvar Hassan Kaleeckal, Ali Nizar Latif, Riyazuddin Mohammad Shaik, Hanan F. Abdul Rahim, Gheyath K. Nasrallah, Mohamed Ghaith Al Kuwari, Hamad Eid Al Romaihi, Adeel A. Butt, Mohamed H. Al-Thani, Abdullatif Al Khal, Roberto Bertollini, Laith J. Abu-Raddad

**Affiliations:** Department of Pathology, Sidra Medicine, Doha, Qatar; Infectious Disease Epidemiology Group, Weill Cornell Medicine-Qatar, Cornell University, Doha, Qatar; World Health Organization Collaborating Centre for Disease Epidemiology Analytics on HIV/AIDS, Sexually Transmitted Infections, and Viral Hepatitis, Weill Cornell Medicine–Qatar, Cornell University, Qatar Foundation – Education City, Doha, Qatar; Biomedical Research Center, Member of QU Health, Qatar University, Doha, Qatar; Department of Biomedical Science, College of Health Sciences, Member of QU Health, Qatar University, Doha, Qatar; Hamad Medical Corporation, Doha, Qatar; Wellcome-Wolfson Institute for Experimental Medicine, Queens University, Belfast, United Kingdom; Department of Mathematics, Statistics, and Physics, Qatar University, Doha, Qatar; Department of Public Health, College of Health Sciences, QU Health, Qatar University, Doha, Qatar; Primary Health Care Corporation, Doha, Qatar; Ministry of Public Health, Doha, Qatar; Department of Population Health Sciences, Weill Cornell Medicine, Cornell University, New York, New York, USA

**Keywords:** SARS-CoV-2, epidemiology, vaccine, variant, infection, dose, case-control

## Abstract

The SARS-CoV-2 Delta (B.1.617.2) variant of concern is expanding globally. Here, we assess real-world effectiveness of the BNT162b2 (Pfizer-BioNTech) and mRNA-1273 (Moderna) vaccines against this variant in the population of Qatar, using a matched test-negative, case-control study design. BNT162b2 effectiveness against any Delta infection, symptomatic or asymptomatic, was 64.2% (95% CI: 38.1-80.1%) ≥14 days after the first dose and before the second dose, but was only 53.5% (95% CI: 43.9-61.4%) ≥14 days after the second dose, in a population in which a large proportion of fully vaccinated persons received their second dose several months earlier. Corresponding effectiveness measures for mRNA-1273 were 79.0% (95% CI: 58.9-90.1%) and 84.8% (95% CI: 75.9-90.8%), respectively. Effectiveness against any severe, critical, or fatal COVID-19 disease due to Delta was 89.7% (95% CI: 61.0-98.1%) for BNT162b2 and 100.0% (95% CI: 41.2-100.0%) for mRNA-1273, ≥14 days after the second dose. Both BNT162b2 and mRNA-1273 are highly effective in preventing Delta hospitalization and death, but less so in preventing infection, particularly for BNT162b2.

## Introduction

The severe acute respiratory syndrome coronavirus 2 (SARS-CoV-2) Delta (B.1.617.2) variant of concern is expanding globally^1^. Appreciable community transmission of this variant was first detected in Qatar toward the end of March, 2021^2-4^. While the incidence of Delta has been increasing, it remained relatively low. Though there has been a recent surge in cases, no evidence for a clear epidemic wave has materialized as of August 10, 2021. In contrast, the introduction of both the Alpha (B.1.1.7) and Beta (B.1.351) variants led to two rapid back-to-back SARS-CoV-2 waves earlier in 2021^2-6^. The high Coronavirus Disease 2019 (COVID-19) vaccine coverage in Qatar at the present time may have slowed Delta transmission. As of August 10, 2021, 87.8% of persons ≥12 years of age have received at least one vaccine dose of either the BNT162b2^7^(Pfizer-BioNTech) vaccine or the mRNA-1273^8^ (Moderna) vaccine, and 73.8% have received both doses^9^. In the present study, we assessed real-world effectiveness of the BNT162b2 and mRNA-1273 vaccines against the Delta variant.

## Results

### Study population

Between December 21, 2020 and July 21, 2021, 906,078 individuals in Qatar received at least one dose of BNT162b2, and 877,354 completed the two-dose regimen. The median date at first dose was April 18, 2021, and the median date at second dose was May 7, 2021. The median time elapsed between the first and second doses was 21 days (interquartile range (IQR): 21-22 days), and 97.7% of individuals received their second dose ≤30 days after the first dose.

During the same time interval, 490,828 individuals received at least one dose of mRNA-1273, and 409,041 completed the two-dose regimen. The median date at first dose was April 18, 2021, and the median date at second dose was May 12, 2021. Median time elapsed between the first and second doses was 28 days (IQR: 28-31 days), and 74.6% of individuals received their second dose ≤30 days after the first dose.

As a consequence of the schedule of arriving vaccine shipments, BNT162b2 vaccine coverage increased incrementally and steadily from December 2020 and up to the present. Meanwhile, the increases in mRNA-1273 vaccine coverage occurred in specific times following arrival of large shipments, and was not appreciable before March of 2021.

Supplementary Data 1-3 provide flowcharts describing the population selection process for investigating and estimating vaccine effectiveness against Delta for the BNT162b2 vaccine (Supplementary Data 1), the mRNA-1273 vaccine (Supplementary Data 2), and any vaccination with either BNT162b2 or mRNA-1273 (Supplementary Data 3). Demographic characteristics of samples used to estimate vaccine effectiveness ≥14 days after the first dose and ≥14 days after the second dose are presented in Tables 1 and 2, respectively. Median age in the study samples was 31-32 years. Qatar has unusually young, diverse demographics, in that only 9% of its residents are ≥50 years of age, and 89% are expatriates from over 150 countries^10,11^.

**Table 1.** Demographic characteristics of cases (PCR-positive for SARS-CoV-2 Delta variant) and controls (PCR-negative) in samples used in the ≥14-days-after-first-dose analysis of vaccine effectiveness of A) BNT162b2, B) mRNA-1273, and C) BNT162b2 or mRNA-1273.

| Study type | A) Effectiveness of BNT162b2 vaccine |  | B) Effectiveness of mRNA-1273 vaccine |  | C) Effectiveness of BNT162b2 or mRNA-1273 vaccines |  |
| --- | --- | --- | --- | --- | --- | --- |
| Characteristics | Cases*<br>(PCR-positive for<br>Delta variant) | Controls*<br>(PCR-negative) | Cases*<br>(PCR-positive for<br>Delta variant) | Controls*<br>(PCR-negative) | Cases*<br>(PCR-positive for<br>Delta variant) | Controls*<br>(PCR-negative) |
| Median age (IQR) — years | 31 (24-37) | 31 (24-37) | 31 (24-37) | 31 (24-37) | 31 (24-37) | 31 (25-38) |
| Age group — no. (%) |  |  |  |  |  |  |
| <20 years | 234 (14.4) | 234 (14.4) | 228 (13.9) | 228 (13.9) | 239 (13.8) | 239 (13.8) |
| 20-29 years | 507 (31.3) | 507 (31.3) | 507 (30.9) | 507 (30.9) | 527 (30.4) | 527 (30.4) |
| 30-39 years | 585 (36.1) | 585 (36.1) | 586 (35.7) | 586 (35.7) | 625 (36.1) | 625 (36.1) |
| 40-49 years | 233 (14.4) | 233 (14.4) | 253 (15.4) | 253 (15.4) | 266 (15.4) | 266 (15.4) |
| 50-59 years | 51 (3.2) | 51 (3.2) | 54 (3.3) | 54 (3.3) | 58 (3.4) | 58 (3.4) |
| 60-69 years | 7 (0.4) | 7 (0.4) | 8 (0.5) | 8 (0.5) | 12 (0.7) | 12 (0.7) |
| 70+ years | 4 (0.3) | 4 (0.3) | 4 (0.2) | 4 (0.2) | 4 (0.2) | 4 (0.2) |
| Sex |  |  |  |  |  |  |
| Male | 1,314 (81.1) | 1,314 (81.1) | 1,336 (81.5) | 1,336 (81.5) | 1,392 (80.4) | 1,392 (80.4) |
| Female | 307 (18.9) | 307 (18.9) | 304 (18.5) | 304 (18.5) | 339 (19.6) | 339 (19.6) |
| Nationality<sup>†</sup> |  |  |  |  |  |  |
| Bangladeshi | 146 (9.0) | 146 (9.0) | 158 (9.6) | 158 (9.6) | 166 (9.6) | 166 (9.6) |
| Egyptian | 10 (0.6) | 10 (0.6) | 10 (0.6) | 10 (0.6) | 13 (0.8) | 13 (0.8) |
| Filipino | 74 (4.6) | 74 (4.6) | 75 (4.6) | 75 (4.6) | 82 (4.7) | 82 (4.7) |
| Indian | 653 (40.3) | 653 (40.3) | 660 (40.2) | 660 (40.2) | 694 (40.1) | 694 (40.1) |
| Nepalese | 160 (9.9) | 160 (9.9) | 167 (10.2) | 167 (10.2) | 169 (9.8) | 169 (9.8) |
| Pakistani | 128 (7.9) | 128 (7.9) | 132 (8.1) | 132 (8.1) | 133 (7.7) | 133 (7.7) |
| Qatari | 226 (13.9) | 226 (13.9) | 215 (13.1) | 215 (13.1) | 233 (13.5) | 233 (13.5) |
| Sri Lankan | 15 (0.9) | 15 (0.9) | 20 (1.2) | 20 (1.2) | 22 (1.3) | 22 (1.3) |
| Sudanese | 13 (0.8) | 13 (0.8) | 12 (0.7) | 12 (0.7) | 14 (0.8) | 14 (0.8) |
| Other nationalities<sup>‡</sup> | 196 (12.1) | 196 (12.1) | 191 (11.7) | 191 (11.7) | 205 (11.8) | 205 (11.8) |
| Reason for PCR testing |  |  |  |  |  |  |
| Clinical suspicion | 472 (29.1) | 472 (29.1) | 482 (29.4) | 482 (29.4) | 518 (29.9) | 518 (29.9) |
| Contact tracing | 146 (9.0) | 146 (9.0) | 148 (9.0) | 148 (9.0) | 157 (9.1) | 157 (9.1) |
| Survey | 179 (11.0) | 179 (11.0) | 178 (10.9) | 178 (10.9) | 193 (11.2) | 193 (11.2) |
| Port of entry | 441 (27.2) | 441 (27.2) | 452 (27.6) | 452 (27.6) | 471 (27.2) | 471 (27.2) |
| Individual request | 259 (16.0) | 259 (16.0) | 260 (15.9) | 260 (15.9) | 265 (15.3) | 265 (15.3) |
| Healthcare routine testing | 40 (2.5) | 40 (2.5) | 38 (2.3) | 38 (2.3) | 42 (2.4) | 42 (2.4) |
| Pre-travel | 84 (5.2) | 84 (5.2) | 82 (5.0) | 82 (5.0) | 85 (4.9) | 85 (4.9) |

\*Cases and controls were matched one-to-one by sex, 10-year age group, nationality, reason for PCR testing, and calendar week of PCR test.
<sup>†</sup>Nationalities were chosen to represent the most populous groups in Qatar.
<sup>‡</sup>These comprise 35 other nationalities in Qatar in sample A, 35 other nationalities in sample B, and 36 other nationalities in sample C.

**Table 2.**
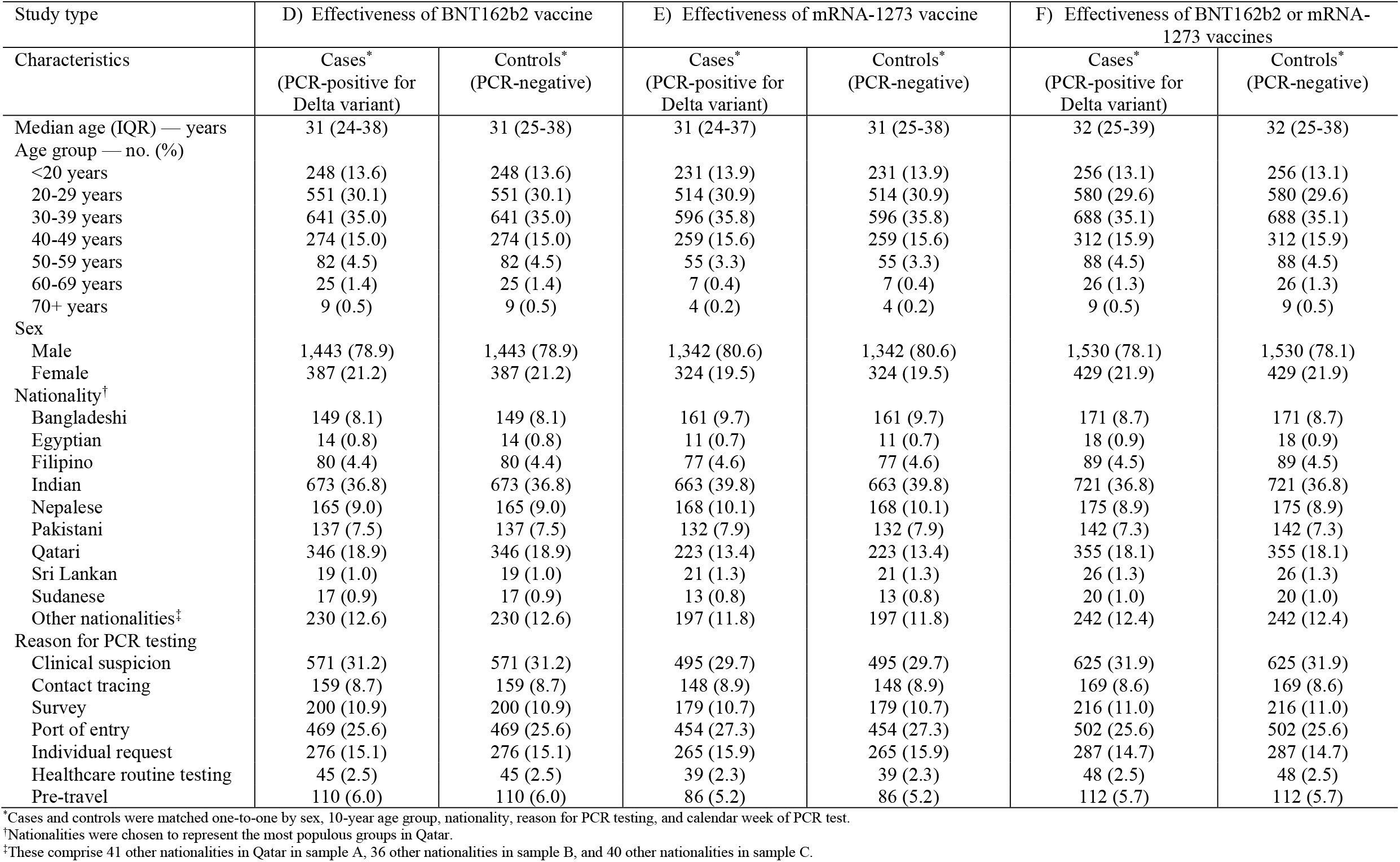
Demographic characteristics of cases (PCR-positive for SARS-CoV-2 Delta variant) and controls (PCR-negative) in samples used in the ≥14-days-after-second-dose analysis of vaccine effectiveness of A) BNT162b2, B) mRNA-1273, and C) BNT162b2 or mRNA-1273.

| Study type | D) Effectiveness of BNT162b2 vaccine |  | E) Effectiveness of mRNA-1273 vaccine |  | F) Effectiveness of BNT162b2 or mRNA-1273 vaccines |  |
| --- | --- | --- | --- | --- | --- | --- |
| Characteristics | Cases*<br>(PCR-positive for<br>Delta variant) | Controls*<br>(PCR-negative) | Cases*<br>(PCR-positive for<br>Delta variant) | Controls*<br>(PCR-negative) | Cases*<br>(PCR-positive for<br>Delta variant) | Controls*<br>(PCR-negative) |
| Median age (IQR) — years | 31 (24-38) | 31 (25-38) | 31 (24-37) | 31 (25-38) | 32 (25-39) | 32 (25-38) |
| Age group — no. (%) |  |  |  |  |  |  |
| <20 years | 248 (13.6) | 248 (13.6) | 231 (13.9) | 231 (13.9) | 256 (13.1) | 256 (13.1) |
| 20-29 years | 551 (30.1) | 551 (30.1) | 514 (30.9) | 514 (30.9) | 580 (29.6) | 580 (29.6) |
| 30-39 years | 641 (35.0) | 641 (35.0) | 596 (35.8) | 596 (35.8) | 688 (35.1) | 688 (35.1) |
| 40-49 years | 274 (15.0) | 274 (15.0) | 259 (15.6) | 259 (15.6) | 312 (15.9) | 312 (15.9) |
| 50-59 years | 82 (4.5) | 82 (4.5) | 55 (3.3) | 55 (3.3) | 88 (4.5) | 88 (4.5) |
| 60-69 years | 25 (1.4) | 25 (1.4) | 7 (0.4) | 7 (0.4) | 26 (1.3) | 26 (1.3) |
| 70+ years | 9 (0.5) | 9 (0.5) | 4 (0.2) | 4 (0.2) | 9 (0.5) | 9 (0.5) |
| Sex |  |  |  |  |  |  |
| Male | 1,443 (78.9) | 1,443 (78.9) | 1,342 (80.6) | 1,342 (80.6) | 1,530 (78.1) | 1,530 (78.1) |
| Female | 387 (21.2) | 387 (21.2) | 324 (19.5) | 324 (19.5) | 429 (21.9) | 429 (21.9) |
| Nationality† |  |  |  |  |  |  |
| Bangladeshi | 149 (8.1) | 149 (8.1) | 161 (9.7) | 161 (9.7) | 171 (8.7) | 171 (8.7) |
| Egyptian | 14 (0.8) | 14 (0.8) | 11 (0.7) | 11 (0.7) | 18 (0.9) | 18 (0.9) |
| Filipino | 80 (4.4) | 80 (4.4) | 77 (4.6) | 77 (4.6) | 89 (4.5) | 89 (4.5) |
| Indian | 673 (36.8) | 673 (36.8) | 663 (39.8) | 663 (39.8) | 721 (36.8) | 721 (36.8) |
| Nepalese | 165 (9.0) | 165 (9.0) | 168 (10.1) | 168 (10.1) | 175 (8.9) | 175 (8.9) |
| Pakistani | 137 (7.5) | 137 (7.5) | 132 (7.9) | 132 (7.9) | 142 (7.3) | 142 (7.3) |
| Qatari | 346 (18.9) | 346 (18.9) | 223 (13.4) | 223 (13.4) | 355 (18.1) | 355 (18.1) |
| Sri Lankan | 19 (1.0) | 19 (1.0) | 21 (1.3) | 21 (1.3) | 26 (1.3) | 26 (1.3) |
| Sudanese | 17 (0.9) | 17 (0.9) | 13 (0.8) | 13 (0.8) | 20 (1.0) | 20 (1.0) |
| Other nationalities‡ | 230 (12.6) | 230 (12.6) | 197 (11.8) | 197 (11.8) | 242 (12.4) | 242 (12.4) |
| Reason for PCR testing |  |  |  |  |  |  |
| Clinical suspicion | 571 (31.2) | 571 (31.2) | 495 (29.7) | 495 (29.7) | 625 (31.9) | 625 (31.9) |
| Contact tracing | 159 (8.7) | 159 (8.7) | 148 (8.9) | 148 (8.9) | 169 (8.6) | 169 (8.6) |
| Survey | 200 (10.9) | 200 (10.9) | 179 (10.7) | 179 (10.7) | 216 (11.0) | 216 (11.0) |
| Port of entry | 469 (25.6) | 469 (25.6) | 454 (27.3) | 454 (27.3) | 502 (25.6) | 502 (25.6) |
| Individual request | 276 (15.1) | 276 (15.1) | 265 (15.9) | 265 (15.9) | 287 (14.7) | 287 (14.7) |
| Healthcare routine testing | 45 (2.5) | 45 (2.5) | 39 (2.3) | 39 (2.3) | 48 (2.5) | 48 (2.5) |
| Pre-travel | 110 (6.0) | 110 (6.0) | 86 (5.2) | 86 (5.2) | 112 (5.7) | 112 (5.7) |

\*Cases and controls were matched one-to-one by sex, 10-year age group, nationality, reason for PCR testing, and calendar week of PCR test.
†Nationalities were chosen to represent the most populous groups in Qatar.
‡These comprise 41 other nationalities in Qatar in sample A, 36 other nationalities in sample B, and 40 other nationalities in sample C.

### Delta vaccine-breakthrough infections

As of the end of the study, July 21, 2021, 54 and 249 Delta breakthrough infections had been recorded among those who received either one or two doses of BNT162b2, respectively, and 27 and 26 breakthrough infections had been recorded among those who received mRNA-1273, respectively.

Also, as of July 21, 2021, 3 and 4 severe Delta COVID-19 disease cases (acute-care hospitalizations^12^; Methods) had been recorded among those who received either one or two doses of BNT162b2, respectively, and 3 severe disease cases had been recorded among those who received only one dose of mRNA-1273. No severe Delta disease cases were recorded among those who received two doses of mRNA-1273.

Meanwhile, 1 critical Delta COVID-19 disease case (ICU-care hospitalization^12^; Methods) had been recorded among those who received two doses of BNT162b2. No critical disease cases were recorded among those who received only one dose of BNT162b2, or among those who received one or two doses of mRNA-1273. No fatal Delta COVID-19 disease cases (COVID-19 deaths^13^; Methods) were recorded among those who received either vaccine.

### Vaccine effectiveness ≥14 days after the first dose

Estimated BNT162b2 effectiveness against infection with Delta, defined as a polymerase chain reaction (PCR)-positive swab with the Delta variant, regardless of the reason for PCR testing or presence of symptoms (Methods), was 64.2% (95% confidence interval (CI): 38.1-80.1%) 14 or more days after the first dose, but before receiving the second dose (Table 3). The corresponding effectiveness for mRNA-1273 was 79.0% (95% CI: 58.9-90.1%), and for vaccination with either BNT162b2 or mRNA-1273, it was 68.9% (95% CI: 52.7-80.1%).

**Table 3.**
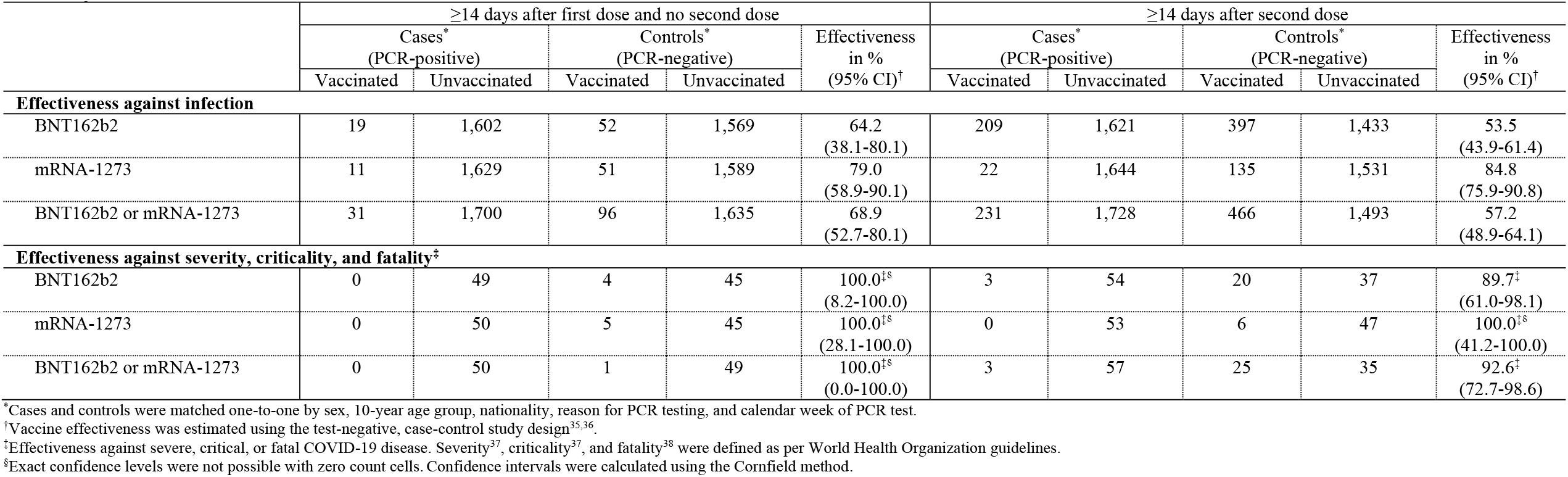
Effectiveness of the BNT162b2 and mRNA-1273 vaccines against the Delta variant ≥14 days after the first dose and ≥14 days after the second dose.

Estimated vaccine effectiveness against any severe^12^, critical^12^, or fatal^13^ COVID-19 disease due to any Delta infection (Methods), 14 or more days after the first dose, but before receiving the second dose, was 100% for BNT162b2, mRNA-1273, and either BNT162b2 or mRNA-1273, but with very wide 95% confidence intervals due to the relatively small number of severe, critical, and fatal disease cases in this analysis (Table 3).

### Vaccine effectiveness ≥14 days after the second dose

Estimated BNT162b2 effectiveness against infection with Delta was 53.5% (95% CI: 43.9- 61.4%) 14 or more days after the second dose (Table 3). The corresponding effectiveness for mRNA-1273 was 84.8% (95% CI: 75.9-90.8%), and for vaccination with either BNT162b2 or mRNA-1273, it was 57.2% (95% CI: 48.9-64.1%).

Estimated BNT162b2 effectiveness against any severe^12^, critical^12^, or fatal^13^ COVID-19 disease due to any Delta infection was 89.7% (95% CI: 61.0-98.1%) 14 or more days after the second dose (Table 3). The corresponding effectiveness for mRNA-1273 was 100.0% (95% CI: 41.2- 100.0%), and for vaccination with either BNT162b2 or mRNA-1273, it was 92.6% (95% CI: 72.7-98.6%).

### Additional analyses

Sensitivity analyses adjusting for sex, age, nationality, reason for PCR testing, and calendar week of PCR test in logistic regression analysis confirmed main analysis results, but with slightly higher estimated effectiveness (Table 4).

**Table 4.**
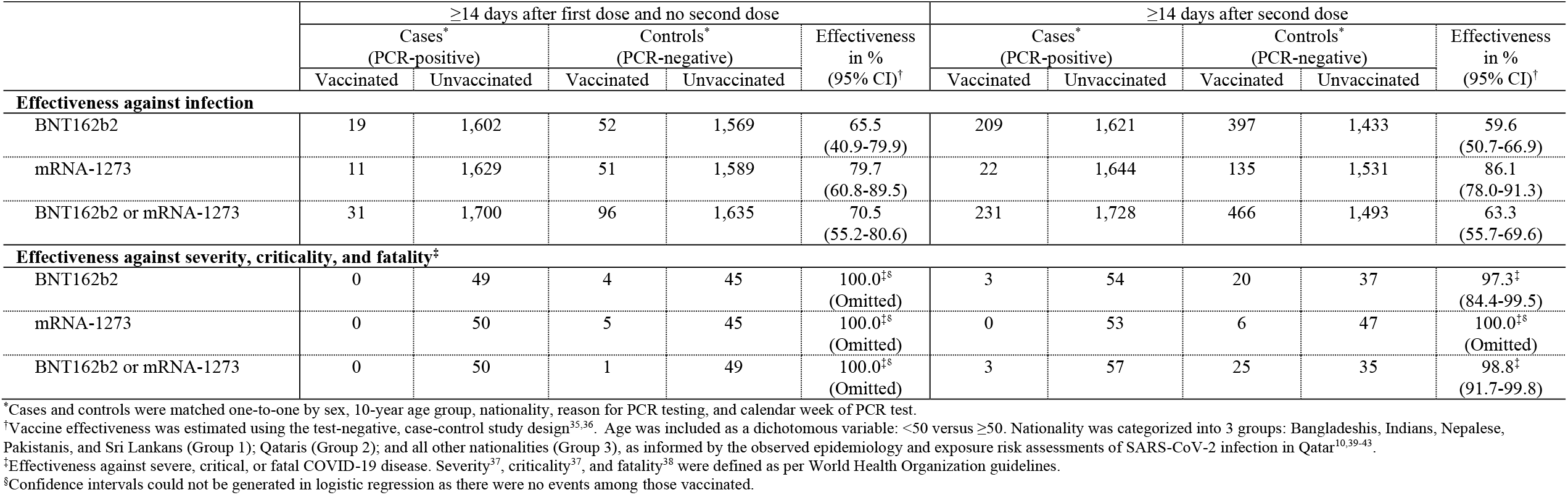
Sensitivity analyses for effectiveness of the BNT162b2 and mRNA-1273 vaccines against the Delta variant ≥14 days after the first dose and ≥14 days after the second dose, adjusting for sex, age, nationality, reason for PCR testing, and calendar week in logistic regression analysis.

An additional analysis estimated BNT162b2 effectiveness against symptomatic Delta infection at 56.1% (95% CI: 41.4-67.2%) 14 or more days after the second dose (Table 5). The corresponding effectiveness for mRNA-1273 was 85.8% (95% CI: 70.6-93.9%), and for vaccination with either BNT162b2 or mRNA-1273, it was 62.0% (95% CI: 50.0-71.1%).

**Table 5.** Effectiveness of the BNT162b2 and mRNA-1273 vaccines against symptomatic and asymptomatic infection with the Delta variant ≥14 days after the first dose and ≥14 days after the second dose.

|  | ≥14 days after first dose and no second dose |  |  |  |  | ≥14 days after second dose |  |  |  |  |
| --- | --- | --- | --- | --- | --- | --- | --- | --- | --- | --- |
|  | Cases*<br>(PCR-positive) |  | Controls*<br>(PCR-negative) |  | Effectiveness<br>in %<br>(95% CI)<sup>†</sup> | Cases*<br>(PCR-positive) |  | Controls*<br>(PCR-negative) |  | Effectiveness<br>in %<br>(95% CI)<sup>†</sup> |
|  | Vaccinated | Unvaccinated | Vaccinated | Unvaccinated |  | Vaccinated | Unvaccinated | Vaccinated | Unvaccinated |  |
| **Effectiveness against symptomatic infection<sup>‡</sup>** |  |  |  |  |  |  |  |  |  |  |
| BNT162b2 | 8 | 464 | 32 | 440 | 76.3<br>(46.7-90.7) | 98 | 473 | 183 | 388 | 56.1<br>(41.4-67.2) |
| mRNA-1273 | 5 | 477 | 33 | 449 | 85.7<br>(62.7-95.7) | 9 | 486 | 57 | 438 | 85.8<br>(70.6-93.9) |
| BNT162b2 or mRNA-1273 | 13 | 505 | 61 | 457 | 80.7<br>(63.9-90.4) | 107 | 518 | 220 | 405 | 62.0<br>(50.0-71.1) |
| **Effectiveness against asymptomatic infection<sup>§</sup>** |  |  |  |  |  |  |  |  |  |  |
| BNT162b2 | 6 | 681 | 8 | 679 | 25.2<br>(0.0-78.7) | 73 | 684 | 108 | 649 | 35.9<br>(11.1-53.9) |
| mRNA-1273 | 3 | 696 | 7 | 692 | 57.4<br>(0.0-92.9) | 7 | 697 | 34 | 670 | 80.2<br>(54.2-92.6) |
| BNT162b2 or mRNA-1273 | 9 | 723 | 16 | 716 | 44.3<br>(0.0-78.4) | 80 | 726 | 125 | 681 | 40.0<br>(18.2-56.1) |

\*Cases and controls were matched one-to-one by sex, 10-year age group, nationality, reason for PCR testing, and calendar week of PCR test.
<sup>†</sup>Vaccine effectiveness was estimated using the test-negative, case-control study design<sup>35,36</sup>.
<sup>‡</sup>A symptomatic infection is defined as a PCR-positive test conducted because of clinical suspicion due to presence of symptoms compatible with a respiratory tract infection.
<sup>§</sup>An asymptomatic infection is defined as a PCR-positive test conducted with no reported presence of symptoms compatible with a respiratory tract infection. That is, the PCR testing was done as part of a survey, for pre-travel requirement, or at the port of entry into the country.

Symptomatic infection was defined as a PCR-positive test conducted because of clinical suspicion due to presence of symptoms compatible with a respiratory tract infection.

An additional analysis estimated BNT162b2 effectiveness against asymptomatic Delta infection at 35.9% (95% CI: 11.1-53.9%) 14 or more days after the second dose (Table 5). The corresponding effectiveness for mRNA-1273 was 80.2% (95% CI: 54.2-92.6%), and for vaccination with either BNT162b2 or mRNA-1273, it was 40.0% (95% CI: 18.2-56.1%). Asymptomatic infection was defined as a PCR-positive test conducted with no reported presence of symptoms compatible with a respiratory tract infection, that is, strictly, the PCR testing was done as only part of a survey, for pre-travel requirement, or at port of entry upon arrival into the country^10,14^.

## Discussion

Both the BNT162b2 and mRNA-1273 vaccines demonstrated robust effectiveness (≥90%) against hospitalization and death due to infection with the Delta variant, confirming recently announced estimates from the United Kingdom (UK)^15^ and Israel^16^. Although there were many breakthrough infections, especially for BNT162b2, severe or critical COVD-19 disease cases among vaccinated persons were rare. Among those fully vaccinated with BNT162b2, there were only 4 severe disease cases and 1 critical disease case due to Delta, while for mRNA-1273, there were only 3 severe disease cases and no critical disease cases. No COVID-19 deaths due to Delta were recorded in a person vaccinated with either vaccine.

BNT162b2 effectiveness against any infection with Delta, symptomatic or asymptomatic, 14 or more days after the first dose, was 64.2%, compared to 46.5%^17^ against Beta (B.1.351) and 65.5%^17^ against Alpha (B.1.1.7), in the same population of Qatar. This result is consistent with existing evidence suggesting that Beta is the most immune-evasive variant of the known variants of concern^18,19^. However, BNT162b2 effectiveness against Delta 14 or more days after the second dose was only 53.5%, compared to 75.0%^5,17^ against Beta and 89.5%^5,17^ against Alpha.

Strikingly, estimated BNT162b2 effectiveness against Delta 14 or more days after the first dose, or 14 or more days after the second dose, were statistically similar.

Emerging evidence suggests significant waning of BNT162b2 effectiveness over time^20-22^. Effectiveness against Beta was estimated at a time when nearly all persons in Qatar were newly vaccinated with BNT162b2^5,17^, but effectiveness against Delta was estimated (in the present study) at a time when a large proportion of those BNT162b2 vaccinated had received their second dose several months earlier. The unexpectedly low effectiveness against Delta among fully vaccinated persons may thus reflect some waning of BNT162b2 protection over time.

This explanation is consistent with the pattern seen in emerging effectiveness estimates against Delta in other countries. Our estimate of 53.5% among fully vaccinated persons is lower than that found in the UK^23,24^ and Canada^25^, where effectiveness was estimated at >75%, but higher than that in Israel at only 39%^16^ and the United States (USA) at 42%^26^. With the delayed second dose in the UK and Canada, most BNT162b2-vaccinated persons received their second dose three months or so more recently than in Israel, USA, and Qatar, where the second dose was administered three weeks after the first dose. The lower effectiveness in Israel, USA, and Qatar may thus reflect waning of vaccine protection for those who received their second dose by end of 2020 or early 2021. Qatar started its mass vaccination campaign with BNT162b2 shortly after that of Israel and the USA.

Another factor may also help to explain this low effectiveness. Public health restrictions have gradually been easing in Qatar over the last few weeks, coinciding with the period during which Delta incidence has been slowly increasing. Restrictions, however, have been eased differently for vaccinated and unvaccinated persons. Many social, work, and travel activities at present require evidence of vaccination (a “health pass”) that is administered through a mandatory mobile app (the Ehteraz app). Vaccinated persons presumably have a higher social contact rate than unvaccinated persons, and may have also reduced their adherence to safety measures, such as protective masks, with the perception of vaccine protection^27-29^. Such risk compensation may be even higher with increasing time since receiving the second dose, leading to progressive normalization of behavior^28-30^. Risk of exposure to the virus could thus be higher among vaccinated persons than among unvaccinated persons, thereby diminishing the observed real-world vaccine effectiveness relative to the actual biological vaccine effectiveness.

mRNA-1273 effectiveness against infection with Delta was higher than that of BNT162b2, consistent with evidence suggesting that mRNA-1273 induces stronger immune response and protection^6,31-33^. mRNA-1273 effectiveness against Delta, 14 or more days after the first dose, was 79.0%, compared to 61.3%^6^ against Beta and 88.1%^6^ against Alpha, in the same population in Qatar. This result is also consistent with Beta’s being the most immune-evasive variant^18,19^. However, mRNA-1273 effectiveness against Delta, 14 or more days after the second dose, was only 84.8%, compared to 96.4%^6^ against Beta and 100%^6^ against Alpha. The same factors that may explain the low BNT162b2 effectiveness against Delta among fully vaccinated persons may also explain the less-than-expected mRNA-1273 effectiveness against Delta among fully vaccinated persons. However, there has been less time for waning of vaccine immunity for mRNA-1273, as this vaccine was incorporated into the national immunization campaign nearly three months after BNT162b2^5,6,17^.

There was evidence for a gradient in vaccine effectiveness by appearance of symptoms, that is, greater protection against more symptomatic or severe infections, as observed earlier for BNT162b2 and mRNA-1273 effectiveness against the Alpha and Beta variants^5,6,17,25^.

This study has limitations. Estimated effectiveness against severe, critical, or fatal COVID-19 disease had wider 95% confidence intervals than that against infection, as a consequence of the smaller number of such cases in the young population of Qatar^10,34^. Data on co-morbid conditions were not available; therefore, they could not be factored explicitly in our analysis. However, adjusting for age may have served as a proxy given that co-morbidities are associated with older age. Furthermore, with the young population of Qatar^10,11^, we anticipate that only a small proportion of the study population may have had serious co-morbid conditions.

Accordingly, our findings may not be generalizable to other settings where the elderly population constitutes a sizable proportion of the population. Effectiveness was assessed using an observational test-negative case-control study design^35,36^, rather than a randomized clinical trial design where cohorts of vaccinated and unvaccinated individuals were being followed up.

However, the cohort study design applied to the same population of Qatar yielded earlier similar findings to the test-negative case-control study design^5,6^, supporting the validity of this standard approach in assessing vaccine effectiveness for respiratory tract infections^35,36^.

In conclusion, both the BNT162b2 and mRNA-1273 vaccines are highly effective in preventing hospitalization and death due to infection with the Delta variant. However, effectiveness against infection was considerably lower than that against serious COVID-19 disease, particularly for the BNT162b2 vaccine. The reasons for the inferior protection against infection remain to be determined, and may not necessarily relate to immune evasion by the Delta variant. The lower effectiveness may reflect some waning of BNT162b2 vaccine protection over time, or higher risk of exposure to the virus among vaccinated persons compared to unvaccinated persons, due to higher social contact rate and less adherence to safety measures.

## Data Availability

The dataset of this study is a property of the Qatar Ministry of Public Health that was provided to the researchers through a restricted-access agreement that prevents sharing the dataset with a third party or publicly. Future access to this dataset can be considered through a direct application for data access to Her Excellency the Minister of Public Health (https://www.moph.gov.qa/english/Pages/default.aspx). Aggregate data are available within the manuscript and its Supplementary information.

## Acknowledgements

We acknowledge the many dedicated individuals at Hamad Medical Corporation, the Ministry of Public Health, the Primary Health Care Corporation, Qatar Biobank, Sidra Medicine, and Weill Cornell Medicine for their diligent efforts and contributions to make this study possible. The authors are grateful for institutional salary support from the Biomedical Research Program and the Biostatistics, Epidemiology, and Biomathematics Research Core, both at Weill Cornell Medicine-Qatar, as well as for institutional salary support provided by the Ministry of Public Health and Hamad Medical Corporation. The authors are also grateful for the Qatar Genome Programme for institutional support for the reagents needed for the viral genome sequencing.

The funders of the study had no role in study design, data collection, data analysis, data interpretation, or writing of the article. Statements made herein are solely the responsibility of the authors.

## Author contributions

PT and MRH conducted the multiplex, RT-qPCR variant screening and viral genome sequencing. HC co-designed the study, performed the statistical analyses, and co-wrote the first draft of the article. LJA conceived and co-designed the study, led the statistical analyses, and co-wrote the first draft of the article. HY, FMB, and HAK conducted viral genome sequencing. All authors contributed to data collection and acquisition, database development, discussion and interpretation of the results, and to the writing of the manuscript. All authors have read and approved the final manuscript.

## Methods

### Data sources, study population, and study design

This study was conducted in the resident population of Qatar. Coronavirus Disease 2019 (COVID-19) laboratory testing, vaccination, clinical infection data, and related demographic details were extracted from the integrated nationwide digital-health information platform that hosts the national, federated SARS-CoV-2 databases. These databases are complete and have captured all SARS-CoV-2-related data since epidemic onset. Nearly all individuals were vaccinated (free of charge) in Qatar and not elsewhere. In rare situations in which an individual received COVID-19 vaccination outside Qatar, the individual’s vaccination details were still recorded in the health system at the port of entry (airport) upon return to Qatar, in order to fulfill national requirements and to benefit from privileges associated with vaccination, such as quarantine exemption.

Vaccine effectiveness was estimated using the test-negative, case-control study design, a standard design for assessing vaccine effectiveness against influenza^35,36^. Key to this design is the control of bias arising from misclassification of infection and differences in health care- seeking behavior between vaccinated and unvaccinated individuals^35,36^. Cases and controls were matched one-to-one by sex, 10-year age group, nationality, reason for SARS-CoV-2 polymerase chain reaction (PCR) testing, and calendar week of PCR test. Matching of cases and controls was performed to control for known differences in the risk of exposure to infection in Qatar^10,40-42^.

Effectiveness was estimated against documented infection (defined as a PCR-positive swab regardless of the reason for PCR testing or presence of symptoms) with the Delta (B.1.617.2) variant, as well as against severe, critical, or fatal COVID-19 disease due to Delta infection. Classification of COVID-19 case severity (acute-care hospitalizations)^12^, criticality (ICU hospitalizations)^12^, and fatality^13^ followed the World Health Organization guidelines, and assessments were made by trained medical personnel using individual chart reviews (see below).

All records of PCR testing for those vaccinated and unvaccinated during the study duration were examined. All persons who received mixed vaccines, or who received a vaccine other than BNT162b2 or mRNA-1273 were excluded. Every case that met the inclusion criteria (a Delta case) and that could be matched to a control was included in the analysis. Both PCR-test outcomes and vaccination status were ascertained at the time of the PCR test. Each person that had a positive PCR test result and hospital admission was subject to an infection severity assessment every three days until discharge or death. Individuals who progressed to COVID-19 disease between the time of the positive PCR test result and the end of the study were classified based on their worst outcome, starting with death^13^, followed by critical disease^12^, and then severe disease^12^ (see below).

The study was approved by the Hamad Medical Corporation and Weill Cornell Medicine-Qatar Institutional Review Boards with waiver of informed consent. The STROBE checklist can be found in Supplementary Table 1.

### COVID-19 severity, criticality, and fatality classification

Severe COVID-19 disease was defined per WHO classification as a SARS-CoV-2 infected person with “oxygen saturation of <90% on room air, and/or respiratory rate of >30 breaths/minute in adults and children >5 years old (or ≥60 breaths/minute in children <2 months old or ≥50 breaths/minute in children 2–11 months old or ≥40 breaths/minute in children 1–5 years old), and/or signs of severe respiratory distress (accessory muscle use and inability to complete full sentences, and, in children, very severe chest wall indrawing, grunting, central cyanosis, or presence of any other general danger signs)”^37^. Detailed WHO criteria for classifying SARS-CoV-2 infection severity can be found in the WHO technical report^37^.

Critical COVID-19 disease was defined per WHO classification as a SARS-CoV-2 infected person with “acute respiratory distress syndrome, sepsis, septic shock, or other conditions that would normally require the provision of life sustaining therapies such as mechanical ventilation (invasive or non-invasive) or vasopressor therapy”^37^. Detailed WHO criteria for classifying SARS-CoV-2 infection criticality can be found in the WHO technical report^37^.

COVID-19 death was defined per WHO classification as “a death resulting from a clinically compatible illness, in a probable or confirmed COVID-19 case, unless there is a clear alternative cause of death that cannot be related to COVID-19 disease (e.g. trauma). There should be no period of complete recovery from COVID-19 between illness and death. A death due to COVID- 19 may not be attributed to another disease (e.g. cancer) and should be counted independently of preexisting conditions that are suspected of triggering a severe course of COVID-19”. Detailed WHO criteria for classifying COVID-19 death can be found in the WHO technical report^38^.

### Classification of infections by variant type

Surveillance for SARS-CoV-2 variants in Qatar is based on viral genome sequencing and multiplex, real-time reverse-transcription PCR (RT-qPCR) variant screening^44^ of random positive clinical samples^2-6^, and complemented by deep sequencing of wastewater samples^2^. The ascertainment of the B.1.617.2 (Delta) cases in this study was based on the results of the weekly RT-qPCR genotyping of the positive clinical samples^2,4^. Between March 23, 2021 and July 21, 2021, RT-qPCR genotyping identified 5,394 (50.6%) B.1.351-like cases, 3,046 (28.6%) B.1.1.7- like cases, 2,175 (20.4%) “other” variant cases, and 41 (0.4%) B.1.375-like and B.1.258-like cases in 10,656 randomly collected SARS-CoV-2-positive specimens^2,4^.

The accuracy of the RT-qPCR genotyping was verified against either Sanger sequencing of the receptor-binding domain (RBD) of SARS-CoV-2 surface glycoprotein (S) gene, or by viral whole-genome sequencing on a Nanopore GridION sequencing device. From 236 random samples (27 B.1.1.7-like, 186 B.1.351/P.1-like, and 23 “other” variants), the PCR genotyping results for B.1.1.7-like, B.1.351/P.1-like, and ‘other’ variants were in 88.8% (23 out of 27), 99.5% (185 out of 186), and 100% (23 out of 23) agreement with the SARS-CoV-2 lineages assigned by sequencing.

Within the “other” variant category, Sanger sequencing and/or Illumina sequencing of the RBD of SARS-CoV-2 spike gene on 450 random samples confirmed that 433 (96.2%) were B.1.617.2 cases, 8 (1.8%) were B.1.617.1 cases, 3 (0.7%) were B.1 cases, 1 (0.2%) was a P.1 case, and 1 (0.2%) was a B.1.617.3 case, with 5 (1.1%) samples failing sequencing^2,4^. Accordingly, a Delta case was proxied as any “other” case identified through the RT-qPCR based variant screening.

### Laboratory methods

Nasopharyngeal and/or oropharyngeal swabs (Huachenyang Technology, China) were collected for PCR testing and placed in Universal Transport Medium (UTM). Aliquots of UTM were: extracted on a QIAsymphony platform (QIAGEN, USA) and tested with real-time reverse- transcription PCR (RT-qPCR) using TaqPath™ COVID-19 Combo Kits (Thermo Fisher Scientific, USA) on an ABI 7500 FAST (ThermoFisher, USA); tested directly on the Cepheid GeneXpert system using the Xpert Xpress SARS-CoV-2 (Cepheid, USA); or loaded directly into a Roche cobas® 6800 system and assayed with a cobas® SARS-CoV-2 Test (Roche, Switzerland). The first assay targets the viral S, N, and ORF1ab gene regions. The second targets the viral N and E-gene regions, and the third targets the ORF1ab and E-gene regions.

All tests were conducted at the HMC Central Laboratory or Sidra Medicine Laboratory, following standardized protocols.

### Statistical analysis

Descriptive statistics (frequency distributions and measures of central tendency) were used to characterize the study samples. The odds ratio, comparing odds of vaccination among cases to that among controls, and its associated 95% confidence interval (CI) were calculated using the exact method. Confidence intervals were not adjusted for multiplicity. Interactions were not investigated. Vaccine effectiveness at different time frames and its associated 95% CI were then calculated by applying the following equation^35,36^:

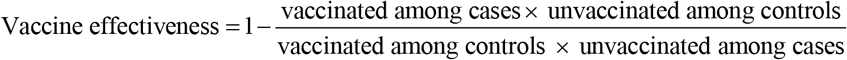

Statistical analyses were conducted in STATA/SE version 17.0^45^.

### Additional analyses

To ensure that vaccine effectiveness estimates were not biased by epidemic phase^35,46^, the gradual roll-out of vaccination during the study^35,46^, and other confounders^47,48^, a sensitivity analysis was conducted by adjusting for the matching factors in logistic regression, that is sex, age, nationality, reason for PCR testing, and calendar week of PCR test (Table 4).

Additional analyses were conducted to estimate vaccine effectiveness against symptomatic infection, defined as a PCR-positive test conducted because of clinical suspicion due to presence of symptoms compatible with a respiratory tract infection, and against asymptomatic infection, defined as a PCR-positive test conducted with no reported presence of symptoms compatible with a respiratory tract infection, that is the PCR testing was done as part of a survey, for pre- travel requirement, or at port of entry into the country.

## Supplementary Material

**Supplementary Data 1.**
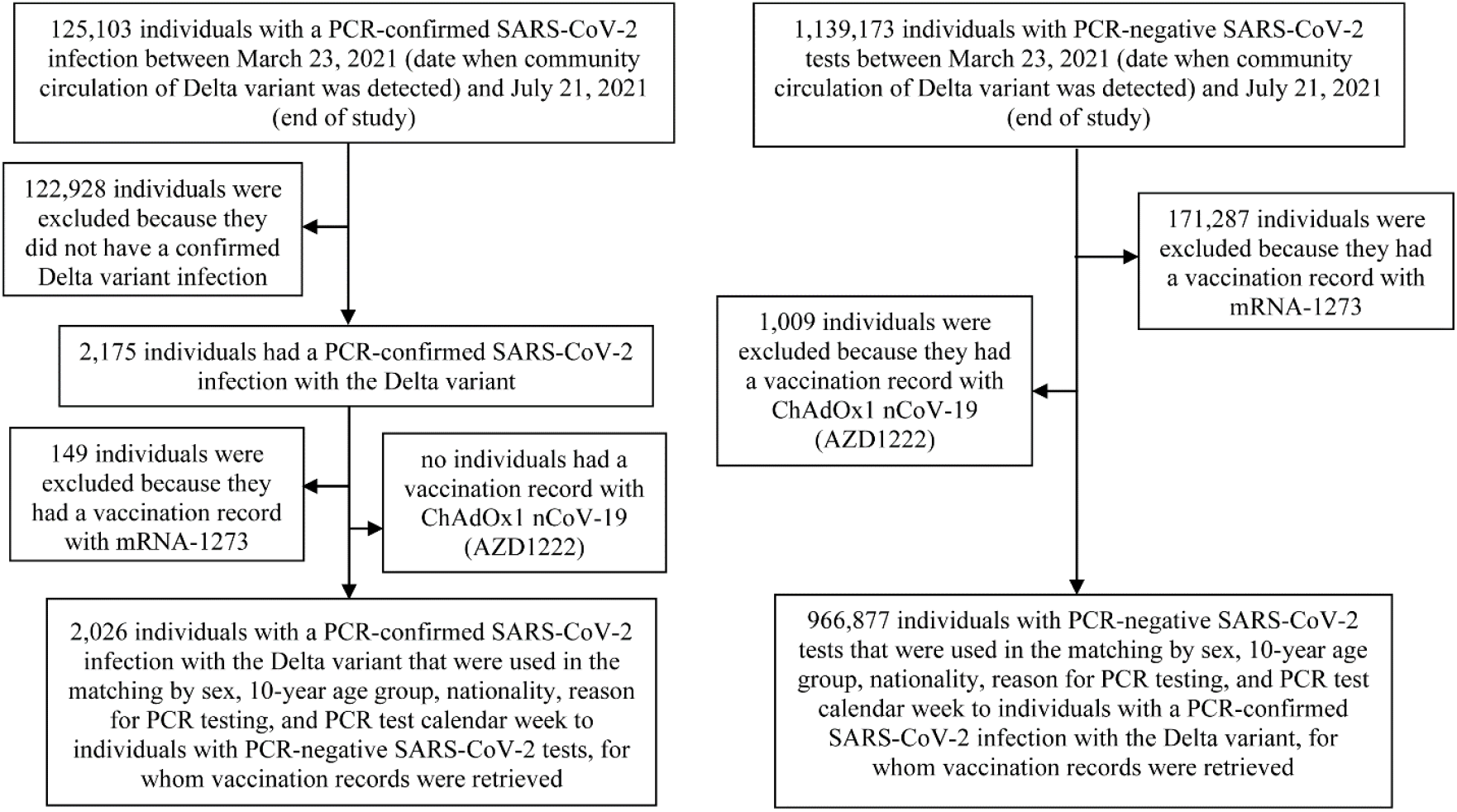
Flowchart describing the population selection process for investigating BNT162b2 vaccine effectiveness against infection with the SARS-CoV-2 Delta variant.

**Supplementary Data 2.**
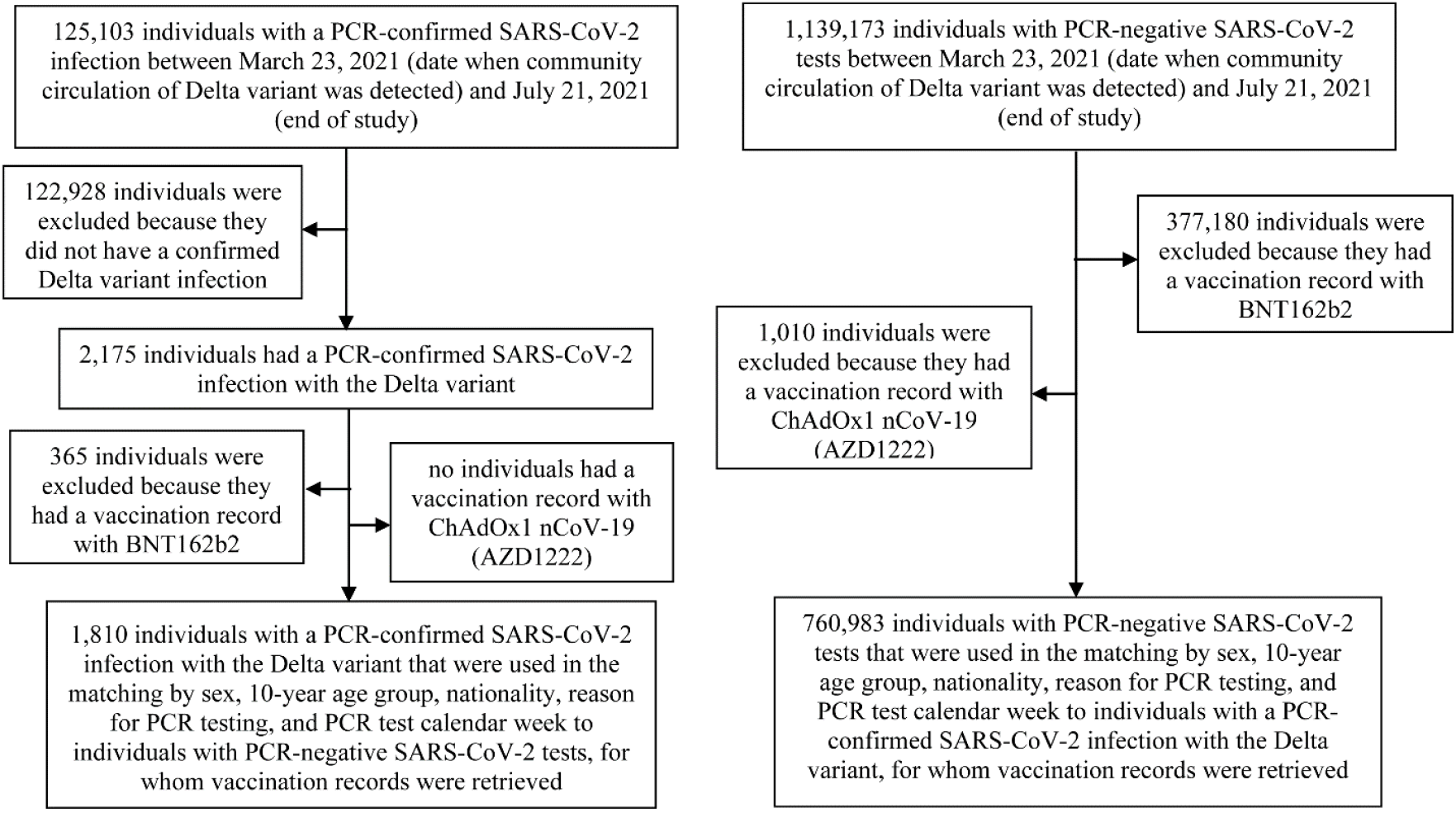
Flowchart describing the population selection process for investigating mRNA-1273 vaccine effectiveness against infection with the SARS-CoV-2 Delta variant.

**Supplementary Data 3.**
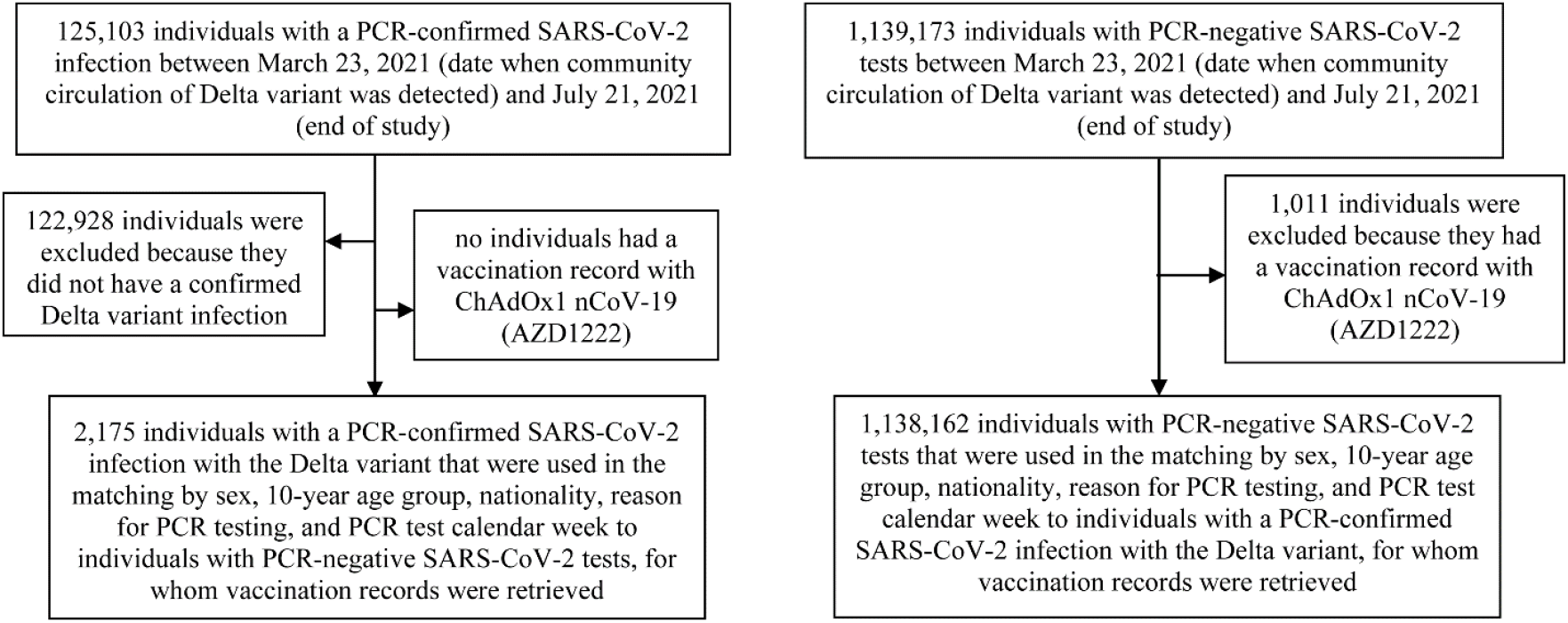
Flowchart describing the population selection process for investigating the BNT162b2 and mRNA-1273 vaccines effectiveness against infection with the SARS-CoV-2 Delta variant.

**Supplementary Table 1.**
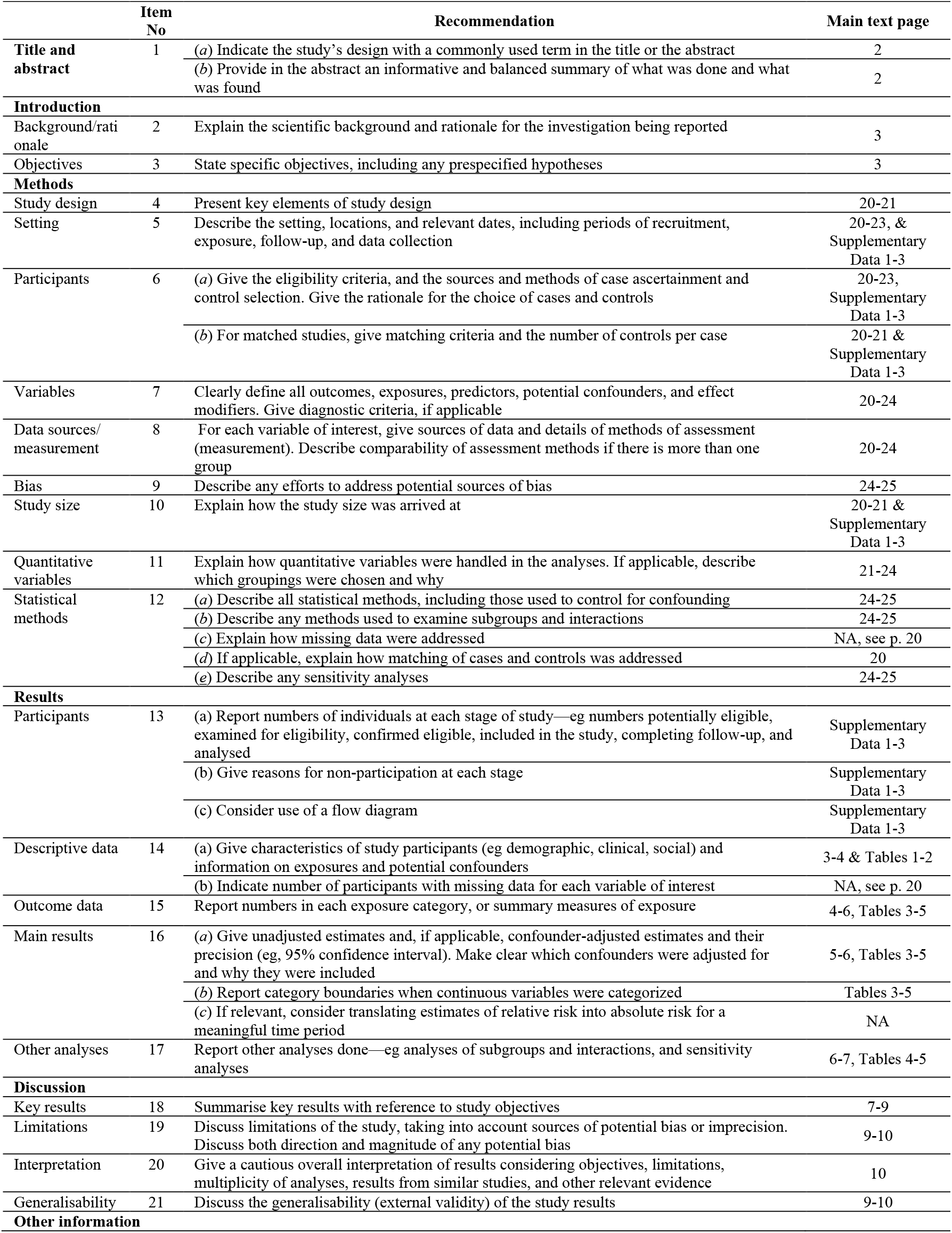
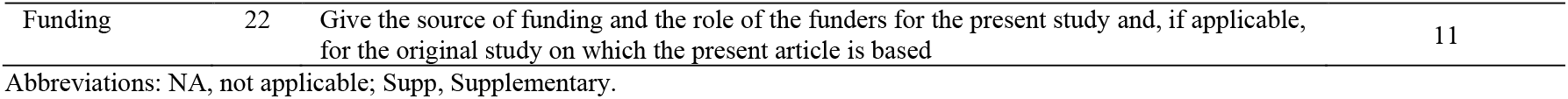
STROBE checklist for case-control studies.

|  | Item No | Recommendation | Main text page |
| --- | --- | --- | --- |
| Title and abstract | 1 | (a) Indicate the study's design with a commonly used term in the title or the abstract | 2 |
|  |  | (b) Provide in the abstract an informative and balanced summary of what was done and what was found | 2 |
| Introduction |  |  |  |
| Background/rationale | 2 | Explain the scientific background and rationale for the investigation being reported | 3 |
| Objectives | 3 | State specific objectives, including any prespecified hypotheses | 3 |
| Methods |  |  |  |
| Study design | 4 | Present key elements of study design | 20-21 |
| Setting | 5 | Describe the setting, locations, and relevant dates, including periods of recruitment, exposure, follow-up, and data collection | 20-23, & Supplementary Data 1-3 |
| Participants | 6 | (a) Give the eligibility criteria, and the sources and methods of case ascertainment and control selection. Give the rationale for the choice of cases and controls | 20-23, Supplementary Data 1-3 |
|  |  | (b) For matched studies, give matching criteria and the number of controls per case | 20-21 & Supplementary Data 1-3 |
| Variables | 7 | Clearly define all outcomes, exposures, predictors, potential confounders, and effect modifiers. Give diagnostic criteria, if applicable | 20-24 |
| Data sources/measurement | 8 | For each variable of interest, give sources of data and details of methods of assessment (measurement). Describe comparability of assessment methods if there is more than one group | 20-24 |
| Bias | 9 | Describe any efforts to address potential sources of bias | 24-25 |
| Study size | 10 | Explain how the study size was arrived at | 20-21 & Supplementary Data 1-3 |
| Quantitative variables | 11 | Explain how quantitative variables were handled in the analyses. If applicable, describe which groupings were chosen and why | 21-24 |
| Statistical methods | 12 | (a) Describe all statistical methods, including those used to control for confounding | 24-25 |
|  |  | (b) Describe any methods used to examine subgroups and interactions | 24-25 |
|  |  | (c) Explain how missing data were addressed | NA, see p. 20 |
|  |  | (d) If applicable, explain how matching of cases and controls was addressed | 20 |
|  |  | (e) Describe any sensitivity analyses | 24-25 |
| Results |  |  |  |
| Participants | 13 | (a) Report numbers of individuals at each stage of study—eg numbers potentially eligible, examined for eligibility, confirmed eligible, included in the study, completing follow-up, and analysed | Supplementary Data 1-3 |
|  |  | (b) Give reasons for non-participation at each stage | Supplementary Data 1-3 |
|  |  | (c) Consider use of a flow diagram | Supplementary Data 1-3 |
| Descriptive data | 14 | (a) Give characteristics of study participants (eg demographic, clinical, social) and information on exposures and potential confounders | 3-4 & Tables 1-2 |
|  |  | (b) Indicate number of participants with missing data for each variable of interest | NA, see p. 20 |
| Outcome data | 15 | Report numbers in each exposure category, or summary measures of exposure | 4-6, Tables 3-5 |
| Main results | 16 | (a) Give unadjusted estimates and, if applicable, confounder-adjusted estimates and their precision (eg, 95% confidence interval). Make clear which confounders were adjusted for and why they were included | 5-6, Tables 3-5 |
|  |  | (b) Report category boundaries when continuous variables were categorized | Tables 3-5 |
|  |  | (c) If relevant, consider translating estimates of relative risk into absolute risk for a meaningful time period | NA |
| Other analyses | 17 | Report other analyses done—eg analyses of subgroups and interactions, and sensitivity analyses | 6-7, Tables 4-5 |
| Discussion |  |  |  |
| Key results | 18 | Summarise key results with reference to study objectives | 7-9 |
| Limitations | 19 | Discuss limitations of the study, taking into account sources of potential bias or imprecision. Discuss both direction and magnitude of any potential bias | 9-10 |
| Interpretation | 20 | Give a cautious overall interpretation of results considering objectives, limitations, multiplicity of analyses, results from similar studies, and other relevant evidence | 10 |
| Generalisability | 21 | Discuss the generalisability (external validity) of the study results | 9-10 |
| Other information |  |  |  |
| Funding | 22 | Give the source of funding and the role of the funders for the present study and, if applicable, for the original study on which the present article is based | 11 |

Abbreviations: NA, not applicable; Supp, Supplementary.

## References

1. World Health Organization. Tracking SARS-CoV-2 variants. Available from: https://www.who.int/en/activities/tracking-SARS-CoV-2-variants/. Accessed on: June 5, 2021. (2021).

2. National Project of Surveillance for Variants of Concern and Viral Genome Sequencing. Qatar viral genome sequencing data. Data on randomly collected samples. https://www.gisaid.org/phylodynamics/global/nextstrain/. (2021).

3. Benslimane, F.M., et al. One year of SARS-CoV-2: Genomic characterization of COVID-19 outbreak in Qatar. medRxiv, 2021.2005.2019.21257433 (2021).

4. Hasan, M.R., et al. Real-Time SARS-CoV-2 Genotyping by High-Throughput Multiplex PCR Reveals the Epidemiology of the Variants of Concern in Qatar. medRxiv, 2021.2007.2018.21260718 (2021).

5. Abu-Raddad, L.J., Chemaitelly, H., Butt, A.A. & National Study Group for Covid-19 Vaccination. Effectiveness of the BNT162b2 Covid-19 Vaccine against the B.1.1.7 and B.1.351 Variants. N Engl J Med (2021).

6. Chemaitelly, H., et al. mRNA-1273 COVID-19 vaccine effectiveness against the B.1.1.7 and B.1.351 variants and severe COVID-19 disease in Qatar. Nat Med (2021).

7. Polack, F.P., et al. Safety and Efficacy of the BNT162b2 mRNA Covid-19 Vaccine. N Engl J Med (2020).

8. Baden, L.R., et al. Efficacy and Safety of the mRNA-1273 SARS-CoV-2 Vaccine. N Engl J Med 384, 403–416 (2021).

9. Ministry of Public Health. National Covid-19 Vaccination Program Data. https://covid19.moph.gov.qa/EN/Pages/Vaccination-Program-Data.aspx. Access date August 2, 2021. (2021).

10. Abu-Raddad, L.J., et al. Characterizing the Qatar advanced-phase SARS-CoV-2 epidemic. Scientific Reports 11, 6233 (2021).

11. Planning and Statistics Authority-State of Qatar. Qatar Monthly Statistics. Available from: https://www.psa.gov.qa/en/pages/default.aspx. Accessed on: May 26, 2020. (2020).

12. World Health Organization. COVID-19 clinical management: living guidance. Available from: https://www.who.int/publications/i/item/WHO-2019-nCoV-clinical-2021-1. Accessed on: May 15, 2021. (2021).

13. World Health Organization. International guidelines for certification and classification (coding) of COVID-19 as cause of death. Available from: https://www.who.int/classifications/icd/Guidelines_Cause_of_Death_COVID-19-20200420-EN.pdf?ua=1. Document Number: WHO/HQ/DDI/DNA/CAT. Accessed on May 15, 2021. (2020).

14. Bertollini, R., et al. Associations of Vaccination and of Prior Infection With Positive PCR Test Results for SARS-CoV-2 in Airline Passengers Arriving in Qatar. JAMA (2021).

15. Stowe, J., et al. Effectiveness of COVID-19 vaccines against hospital admission with the Delta (B.1.617.2) variant. https://khub.net/web/phe-national/public-library/-/document_library/v2WsRK3ZlEig/view_file/479607329?_com_liferay_document_library_web_portlet_DLPortlet_INSTANCE_v2WsRK3ZlEig_redirect=https%3A%2F%2Fkhub.net%3A443%2Fweb%2Fphe-national%2Fpublic-library%2F-%. (2021).

16. Israel’s Ministry of Health. COVID-19 vaccine effectiveness against the Delta variant. Israel’s Ministry of Health report. https://www.gov.il/BlobFolder/reports/vaccine-efficacy-safety-follow-up-committee/he/files_publications_corona_two-dose-vaccination-data.pdf. (2021).

17. Abu-Raddad, L.J., et al. Pfizer-BioNTech mRNA BNT162b2 Covid-19 vaccine protection against variants of concern after one versus two doses. J Travel Med (2021).

18. Wang, P., et al. Antibody resistance of SARS-CoV-2 variants B.1.351 and B.1.1.7. Nature (2021).

19. Planas, D., et al. Sensitivity of infectious SARS-CoV-2 B.1.1.7 and B.1.351 variants to neutralizing antibodies. Nat Med (2021).

20. Israel, A., et al. Elapsed time since BNT162b2 vaccine and risk of SARS-CoV-2 infection in a large cohort. medRxiv, 2021.2008.2003.21261496 (2021).

21. Thomas, S.J., et al. Six Month Safety and Efficacy of the BNT162b2 mRNA COVID-19 Vaccine. medRxiv, 2021.2007.2028.21261159 (2021).

22. Mizrahi, B., et al. Correlation of SARS-CoV-2 Breakthrough Infections to Time-from-vaccine; Preliminary Study. medRxiv, 2021.2007.2029.21261317 (2021).

23. Lopez Bernal, J., et al. Effectiveness of Covid-19 Vaccines against the B.1.617.2 (Delta) Variant. N Engl J Med (2021).

24. Sheikh, A., McMenamin, J., Taylor, B. & Robertson, C. SARS-CoV-2 Delta VOC in Scotland: demographics, risk of hospital admission, and vaccine effectiveness. The Lancet 397, 2461–2462 (2021).

25. Nasreen, S., et al. Effectiveness of COVID-19 vaccines against variants of concern, Canada. medRxiv, 2021.2006.2028.21259420 (2021).

26. Puranik, A., et al. Comparison of two highly-effective mRNA vaccines for COVID-19 during periods of Alpha and Delta variant prevalence. medRxiv, 2021.2008.2006.21261707 (2021).

27. Makhoul M., et al. Epidemiological impact of SARS-CoV-2 vaccination: Mathematical modeling analyses. Vaccines 8(2020).

28. Usherwood, T., LaJoie, Z. & Srivastava, V. A model and predictions for COVID-19 considering population behavior and vaccination. Scientific Reports 11, 12051 (2021).

29. Andersson, O., Campos-Mercade, P., Meier, A.N. & Wengström, E. Anticipation of COVID-19 Vaccines Reduces Social Distancing. https://papers.ssrn.com/sol3/papers.cfm?abstract_id=3765329. SSRN (2021).

30. Cassell, M.M., Halperin, D.T., Shelton, J.D. & Stanton, D. Risk compensation: the Achilles’ heel of innovations in HIV prevention? BMJ 332, 605–607 (2006).

31. Abu-Raddad, L.J., et al. Effect of vaccination and of prior infection on infectiousness of vaccine breakthrough infections and reinfections. medRxiv, 2021.2007.2028.21261086 (2021).

32. Abu-Raddad, L.J., et al. Protection afforded by the BNT162b2 and mRNA-1273 COVID-19 vaccines in fully vaccinated cohorts with and without prior infection. medRxiv, 2021.2007.2025.21261093 (2021).

33. Khoury, D.S., et al. Neutralizing antibody levels are highly predictive of immune protection from symptomatic SARS-CoV-2 infection. Nat Med 27, 1205–1211 (2021).

34. Seedat, S., et al. SARS-CoV-2 infection hospitalization, severity, criticality, and fatality rates. medRxiv 2020.11.29.20240416 (2020).

35. Jackson, M.L. & Nelson, J.C. The test-negative design for estimating influenza vaccine effectiveness. Vaccine 31, 2165–2168 (2013).

36. Verani, J.R., et al. Case-control vaccine effectiveness studies: Preparation, design, and enrollment of cases and controls. Vaccine 35, 3295–3302 (2017).

37. World Health Organization. COVID-19 clinical management: living guidance. Available from: https://www.who.int/publications/i/item/WHO-2019-nCoV-clinical-2021-1. Accessed on: May 15 2021. (2021).

38. World Health Organization. International guidelines for certification and classification (coding) of COVID-19 as cause of death. Available from: https://www.who.int/classifications/icd/Guidelines_Cause_of_Death_COVID-19-20200420-EN.pdf?ua=1. Document Number: WHO/HQ/DDI/DNA/CAT. Accessed on May 31, 2021. (2021).

39. Ayoub, H.H., et al. Mathematical modeling of the SARS-CoV-2 epidemic in Qatar and its impact on the national response to COVID-19. J Glob Health 11, 05005 (2021).

40. Coyle, P.V., et al. SARS-CoV-2 seroprevalence in the urban population of Qatar: An analysis of antibody testing on a sample of 112,941 individuals. iScience, 102646 (2021).

41. Al-Thani, M.H., et al. SARS-CoV-2 infection is at herd immunity in the majority segment of the population of Qatar. Open Forum Infectious Diseases (2021).

42. Jeremijenko, A., et al. Herd Immunity against Severe Acute Respiratory Syndrome Coronavirus 2 Infection in 10 Communities, Qatar. Emerg Infect Dis 27, 1343–1352 (2021).

43. Abu-Raddad, L.J., et al. COVID-19 risk score as a public health tool to guide targeted testing: A demonstration study in Qatar. medRxiv, 2021.2003.2006.21252601 (2021).

44. Vogels, C., Fauver, J. & Grubaugh, N. Multiplexed RT-qPCR to screen for SARS-COV-2 B.1.1.7, B.1.351, and P.1 variants of concern V.3. dx.doi.org/10.17504/protocols.io.br9vm966. (2021).

45. StataCorp. Stata Statistical Software: Release 17. College Station, TX: StataCorp LLC. (2021).

46. Jacoby, P. & Kelly, H. Is it necessary to adjust for calendar time in a test negative design?: Responding to: Jackson ML, Nelson JC. The test negative design for estimating influenza vaccine effectiveness. Vaccine 2013;31(April (17)):2165-8. Vaccine 32, 2942 (2014).

47. Pearce, N. Analysis of matched case-control studies. BMJ 352, i969 (2016).

48. Rothman, K.J., Greenland, S. & Lash, T.L. Modern epidemiology, (Wolters Kluwer Health/Lippincott Williams & Wilkins, Philadelphia, 2008).

